# Intra-urban inequalities in modern family planning use in Uganda’s urban settings: the role of place of residence, socio-economic, family, and individual factors

**DOI:** 10.1101/2023.04.11.23288416

**Authors:** Rornald Muhumuza Kananura, Catherine Birabwa, Ronald Wasswa, Jacquellyn Nambi Ssanyu, Kharim Mwebaza Muluya, Sara Namutamba, Moses Kyangwa, Felix Kizito, Othman Kakaire, Richard Mugahi, Peter Waiswa

**Affiliations:** Department of Health Planning and Management, Makerere University School of Public health, Kampala, Uganda; Center of excellence for Maternal and Newborn Health, Makerere University School of Public Health, Kampala, Uganda; Busoga Health Forum, Jinja, Uganda; Iganga Municipality Local Government, Iganga, Uganda; Department of Public Health, Faculty of Health Sciences, Islamic University in Uganda; Department of Obstetrics and Gynecology, Makerere University School of Medicine, College of Health Sciences, Kampala, Uganda; Ministry of Health, Kampala, Uganda

**Keywords:** Modern family planning use, inequality, urban, cities, Uganda, sub-Sharan Africa

## Abstract

Evidence of how intra-urban inequalities could steer the use of modern family planning (mFP) services remains elusive. In this study, we examined the role of residence, socio-economic, family, and individual factors in shaping access to mFP use in Iganga Municipality and Jinja city, in central eastern Uganda.

We used cross-sectional household survey data that were collected between November-December 2021 from 1023 women aged 15-49 years. We used logistic regression to assess the factors associated with mFP use and Stata user written command – *iop*, to assess the inequality in mFP due to different factors. We considered unfair circumstances as socio-economic status (wealth quartile, education level, and working level), place of residence, age, religion affiliation, and authority. The time of sexual intercourse was considered as fair circumstance.

Overall mFP use was estimated at 48.8%, with close to 60% using long-term acting reversible methods. Overall, 24% of all heterogeneity in modern FP use was due to the observed circumstances and 18% was due to differential in unfair circumstances. An increase in age was inversely associated with mFP use [adjusted Odds Ratio (aOR) [95% Confidence Interval (95%CI)] =0.976[0.966-0.986]), while an increase in parity was positively associated with mFP use (aOR [95%CI]=1.404[1.249-1.578]). Compared to mainland non-slum and landing site residents, mainland slum residents were two-fold (aOR[95%CI]=2.065[1.735-2.458]) and three-fold (aOR[95%CI]=2.631[1.96-3.531]) more likely to use mFP, respectively. Whereas the odds of using mFP increased with the wealth status (Middle: aOR[95%CI]=1.832[1.52-2.209] and Better: aOR[95%CI]=5.276[4.082-6.819]), an interaction between the place of residence and wealth index showed that wealth index mattered only in non-slum mainland areas. Women with secondary or higher level of education and whose decisions to use mFP were independent of other authorities were more likely to use mFP. Lastly, there were region affiliation and type of work differential in the use of mFP.

In conclusion, about one-fifth of all heterogeneity in mFP use was due to differentials in unfair circumstances. The findings highlight the need for intervention that are tailored to the different groups of urban residents. For instance, the package of interventions should consider the places of work and places of residences regardless of socioeconomic status.

## Introduction

Universal access to sexual and reproductive health has been recognized as a component of sustainable development [1, 2]. Empowering women in deciding the number and when to have children contributes substantially to reducing maternal and child mortality and promoting economic growth [3–5]. Available evidence has indicated how modern family planning is critical in reducing the world’s total fertility rate and its consequences including climate change [2]. While the demand for family planning has increased worldwide the unmet need for family planning remains high in developing countries, particularly in poorest countries with the highest fertility rates, and lower levels of women empowerment [6–8]. Within-countries disparities are also observed, with lower levels of coverage among poorer, uneducated, rural, and younger women.

To achieve United Nations’ Sustainable Development Goals 3 and 5 aim of achieving universal access to sexual and reproductive health, without leaving anybody behind, there is a need to understand the inequality in access to family planning. While it has been assumed that urban dwellers have better access to health services, the urban population experience multiple factors that inhibit them from accessing appropriate family planning services [9, 10]. On the one hand, more often the urban healthcare system is dominated by the private health providers [11], who usually provide services at exorbitant prices and usually of poor quality. As such, the poor may not have enough money to purchase appropriate family planning products. On the other hand, the urban population are engaged in employment, most of which are inflexible to allow them access health services because often, they work throughout the day and weekends without leave days [9]. Such kind of employment coupled with other competing family duties may hinder women from accessing appropriate family planning services [9].

Health inequity exists when people are unfairly deprived of the resources that are necessary to prevent them from undesirable conditions [9, 12]. The WHO commission on social determinants of health considers unfair differences within and between groups as social injustice [13]. It is only through the equity lens that certain population segments can be observed if they are being deprived of the family planning resources needed to avoid unwanted pregnancies [9, 12]. Additionally, understanding inequity in family planning services help the policy makers to understands the capacity of the existing public health system in meeting the needs of the most vulnerable individuals [14]. The COVID-19 pandemic has exacerbated the inequities in access to family planning with women in lower socio-economic class areas experiencing inadequacies in access services [15]. This is because women in lower socioeconomic class are usually powerless to make a safe and informed decision.

With the rapid urbanization in developing countries, efforts to achieve universal access to family planning services require an approach that goes beyond urban-rural inequalities to understanding of intra-urban disparities in family planning coverage and the identification of population segments that are being left behind. While utilization of modern family planning is increasing in Uganda, evidence on segments of women within the urban areas remains elusive. Understanding the family planning disparities in urban space is critical in designing facility planning promotion interventions for different segments of the population. In this study we examine the role of socio-economic, family, and individual factors in shaping to the utilization of modern family planning use in Iganga Municipality and Jinja city, in central eastern Uganda. We considered the socio-economic, place of residence, religion affiliation, and demographic factors as unfair circumstances. We argue that women should be able to use family planning services regardless of their socio-economic status, place of residence, religion affiliation and age.

## Methods

### Study design, setting and population.

This was a quantitative cross-sectional household survey that was carried out in Iganga Municipality and Jinja City in central eastern Uganda (Busoga region) from November-December 2021. According to the 2016 Uganda Demographic and Health Survey, the number of children per woman in Busoga region is 6.1 which is higher than the national estimate of 5.4 [16]. In addition, the percentage of currently married women aged 15-49 with unmet need for family planning in Busoga region is estimated at 22% and 15% for unmet need for spacing and limiting respectively. Additionally, unmet need among sexually active unmarried women in the region is estimated at 27% [16].

According to Jinja City and Iganga Municipality statistical abstract reports, during the day 40% of the population is non-resident, who reside in the bordering villages. Therefore, to cater for such differences, we used a list of all the households and workplaces within the selected urban centers. The places of work were saloons, bars/restaurants and markets.

### Sample size and sampling procedure

We applied lot quality assurance sampling (LSQA) data collection methodology. At the first stage of sampling, municipality divisions in the two districts were randomly selected (2 in Iganga and 3 in Jinja). We then randomly selected 5 parishes in each division as LQAS supervision areas, where a fixed sample size of at least 20 households was allocated. Each parish had a sampling frame consisting of all villages with their respective population. This was followed by a random selection of 50% of the villages in each parish using a table of random numbers. The number of interviews in each parish was divided across selected villages.

In the process, a total of 1023 women aged 15-49 years were successful enrolled for the study. The inclusion criteria were women of reproductive age group (15-49 years of age) who were residing and or working in the study area. Of these, 206 women were interviewed from their workplace and therefore information specifically on the household characteristics such as household assets, housing structure, as well as water, hygiene and sanitation were not captured. The study not only excluded women who refused to consent but also those who had severe illness at the time of the survey. Details on the study design and sampling are indicated in the study protocol [17].

### Data collection

Data were collected using face-to-face interview structured questionnaire for each of the sampled households. Twenty experienced and qualified research assistants (6 men and 14 women) were recruited and trained intensively for 7 days with the training covering the data collection tools as well as research and field work ethics. We collected information on woman’s socio-demographic characteristics, marriage, and sexual activity, pre and postnatal care, contraception, child health, fertility (birth and pregnancy history) as well as working environment. The questionnaire was prepared in English and then translated to Lusoga language used in the study areas.

The tool was then uploaded on android tablets using Kobo collect and data collected electronically by the trained research assistants. The tool also had inbuilt validation checks for providing appropriate questions for specific participants and for providing warning messages in case of inconsistence. Prior to data collection, a pretest survey was conducted in villages that were not part of the sampled areas and identified problems were corrected before the actual survey was done. Household and workplace geo-coordinates for all interviews were also taken. Four supervisors were selected and the principal investigator supervised the entire data collection process.

### Study variables and their measurements

Modern Family Planning (mFP) use was the dependent variable. This was defined as the use by a woman or her partner of at least one of the following methods: male or female sterilization, injectable, intrauterine devices, contraceptive pills, implants, female or male condoms, standard-days method, lactational amenorrhea, and emergency contraception. We generated mFP use as a binary outcome which was coded 1 if a woman was currently using any of the above methods at the time of the survey and 0 if a woman was either using a traditional method or not using any method.

The independent variables included woman’s age, education level (no education/primary, secondary/higher), marital status (unmarried, married), and place of residence (landing site slum, mainland slum, and other residential areas such as those for elites and business people). Other variables were working status (Not working, work throughout the year, work seasonally/part of the year, and occasionally), religion (Catholic, Anglicans, Muslims, and Pentecostal) and parity (0, 1, 2, 3, 4, and 5+). Women were further asked to state whether religion is very important with the possible responses of no/yes.

We also included wealth index, which was generated using tetrachoric factor analyses. We included items that are known to determine the household’s wealth status. The items included asset ownership such as television, bicycle, car, and land; housing structure; and household hygiene and access to water sources. From this, wealth tertile was created (poorer, middle, and less poor) were generated with the first tertile representing 33.3% of poor households and the last tertile corresponding to 33.3% less poor households. Family planning decision making authority was another variable where women were asked to state a person who usually decides for them when to use family planning methods and the options were: woman alone, husband, jointly with partner, and other people. The survey also asked women about their sexual activity and the time they last had sex with any man. This was a basis of generating the variable of sexual activity with categories of last month, between 2-5 months, between 6-12 months, and 1 year and above.

### Data analysis

We started with a descriptive summary of all study variables using weighted and unweighted proportions. We then provided the inequalities in the utilization of different family planning services by respondent’s characteristics. Family planning methods were categorized into three (short term, long term reversible contraceptives, and permanent methods). This was followed by running a bivariate and multivariate logistic model to assess the factors associated with modern family planning use among women in the urban setting. Variables with a p-value less than 0.2 at bivariate level were considered for further analysis and fitted in to multivariate binary logistic regression model. A p**-**value < 0.05 in multivariable analysis was considered statistically significant. Using *iop* Stata user written command, we assessed the heterogeneity in utilization of modern family planning due to the factors considered in the multivariate model.

### Ethical considerations

This study was approved by Makerere University School of Public Health Research and Ethics Committee (Ref: SPH-2021-146) and Uganda National Council for Science and Technology (Ref: HS1826ES). Before data collection, we also sought administrative clearance from the leaders of Jinja city and Iganga municipality. Written informed consent was also sought from all participants and anonymity was ensured of all the data collected. All data files were stored on password protected computers to ensure privacy and confidentiality for the respondents.

#### Informed consent process

Written informed consent to participate in the research was obtained from all respondents.

#### Data protection

The data used in this study is part of a lager family planning baseline data that is adequately protected with access limited to only data managers.

## Results

Table 1 shows the distribution of women by selected characteristics. Overall, a total of 1023 women participated in the study. Twenty-eight percent of women were aged 20-24 whilst 26% and 18% were 25-29 and 35-49 years respectively. The study also shows that majority of the surveyed women were staying within slums with about 42% in the mainland slum and 24% on the slums along landing sites. Further, majority of women were married (69%), had secondary/higher education (63%), had sex within the last 2-5 months (64%), and had at least one child (73%). Also, three in ten women were in the third quartile (31%), and either belonged to Muslim (33%) or Anglican (29%) faith. In addition, more than half of women (62%) also acknowledged religion to be so important their daily life. In addition, half of women were using modern family planning (49%), were working throughout the year (49%) and their decision to use FP was decided on by other people (50%).

**Table 1:** Distribution of women by selected characteristics

| Variable | %<br>(Unweighted) | %<br>(Weighted) | Sample<br>(N=1,023) |
| --- | --- | --- | --- |
| **Woman's age** |  |  |  |
| 15-19 | 11.5 | 11.6 | 118 |
| 20-24 | 29.5 | 27.5 | 302 |
| 25-29 | 25.5 | 26.4 | 261 |
| 30-34 | 15.0 | 16.4 | 153 |
| 35-49 | 18.5 | 18.1 | 189 |
| **Place of residence** |  |  |  |
| Landing site slum | 14.2 | 23.5 | 145 |
| Mainland slum | 50.4 | 42.4 | 516 |
| Other residential places | 35.4 | 34.1 | 362 |
| **Marital status** |  |  |  |
| Unmarried | 29.5 | 31.2 | 302 |
| Married | 70.5 | 68.8 | 721 |
| **Education level** |  |  |  |
| No education/Primary | 35.1 | 37.5 | 359 |
| Secondary/higher | 64.9 | 62.5 | 664 |
| **Wealth quintile** |  |  |  |
| 33.3% of the lowest class | 21.5 | 17.8 | 220 |
| 33.3% of the middle class | 25.4 | 27.0 | 260 |
| 33.3% of a better class | 32.9 | 30.5 | 337 |
| Missing** | 20.2 | 24.7 | 206 |
| **FP decision making authority** |  |  |  |
| Woman alone | 6.3 | 5.8 | 64 |
| Husband | 37.9 | 38.5 | 388 |
| Jointly with husband | 7.1 | 6.2 | 73 |
| Other people | 48.7 | 49.5 | 498 |
| **Working status** |  |  |  |
| Not working | 33.4 | 34.0 | 342 |
| Work throughout the year | 48.2 | 48.5 | 493 |
| Work seasonally/Part of the Year | 11.8 | 11.4 | 121 |
| Occasionally | 6.6 | 6.1 | 67 |
| **Religion** |  |  |  |
| Catholic | 17.1 | 17.6 | 175 |
| Muslim | 34.4 | 32.9 | 352 |
| Anglican | 27.5 | 28.5 | 281 |
| Pentecostal | 21.0 | 21.0 | 215 |
| **Religion not very important** |  |  |  |
| No | 57.7 | 61.6 | 590 |
| Yes | 42.1 | 38.3 | 431 |
| Missing | 0.2 | 0.1 | 2 |
| **Sexual activity** |  |  |  |
| Last month | 18.9 | 19.3 | 193 |
| Between 2-5 months | 66.6 | 64.0 | 682 |
| Between 6-12 month | 5.4 | 5.9 | 55 |
| 1 years and above | 6.3 | 7.6 | 64 |
| Missing | 2.8 | 3.2 | 29 |
| **Parity** |  |  |  |
| 0 | 26.4 | 27.2 | 270 |
| 1 | 23.9 | 22.6 | 245 |
| 2 | 19.1 | 16.6 | 195 |
| 3 | 12.5 | 14.7 | 128 |
| 4 | 7.8 | 8.7 | 80 |
| 5+ | 10.3 | 10.2 | 105 |
| **Modern family planning use** |  |  |  |
| No | 50.5 | 51.2 | 517 |
| Yes | 49.5 | 48.8 | 506 |

*\*\* We did not capture information on household characteristics for women whom we interviewed at their workplace*

Table 2 shows the proportion of women using mFP according to their characteristics. In the results, mFP use was lower among women aged 35-49 years (17%), those staying in the slums along landing sites (23%), the unmarried (26%), those who could independently make decision on FP (4%), and those who are not sexually active (2%). Regarding education, mFP was higher among those with secondary/higher education (61%). Modern family planning use also increases with household wealth. For instance, 20%, 39%, and 41% of women in the first, second, and third wealth quintile were respectively using mFP. Still, modern FP use was higher among women with zero parity compared to those with at least 5 children (27% versus 13%). In relation to religion, mFP use was almost the same among Muslims (33%) and Anglican (30%); as well as Catholics (18%) and Pentecostal (19%).

**Table 2:** Distribution of women by type of family planning service

| Variable | Short term method (%) | Long term reversible (%) | Permanent method (%) | Any modern method (%) |
| --- | --- | --- | --- | --- |
| **Woman's age** |  |  |  |  |
| 15-19 | 74.0 | 26.0 | 0.0 | 12.4 |
| 20-24 | 45.9 | 54.1 | 0.0 | 27.1 |
| 25-29 | 31.0 | 69.0 | 0.0 | 23.7 |
| 30-34 | 19.0 | 77.6 | 3.3 | 19.9 |
| 35-49 | 46.2 | 52.6 | 1.2 | 17.0 |
| **Place of residence** |  |  |  |  |
| Landing site slum | 41.2 | 55.9 | 2.9 | 22.6 |
| Mainland slum | 37.1 | 62.5 | 0.4 | 43.3 |
| Other residential places | 44.5 | 55.3 | 0.2 | 34.1 |
| **Marital status** |  |  |  |  |
| Unmarried | 70.0 | 30.0 | 0.0 | 26.4 |
| Married | 30.0 | 68.8 | 1.2 | 73.6 |
| **Education level** |  |  |  |  |
| No education/Primary | 30.9 | 67.0 | 2.2 | 39.5 |
| Secondary/higher | 46.9 | 53.1 | 0.0 | 60.5 |
| **Wealth index** |  |  |  |  |
| 33.3% of the lowest class | 46.5 | 47.8 | 5.7 | 19.8 |
| 33.3% of the middle class | 37.5 | 62.5 | 0.0 | 38.8 |
| 33.3% of a better class | 38.4 | 61.6 | 0.0 | 41.4 |
| **FP decision making authority** |  |  |  |  |
| Woman alone | 70.2 | 29.8 | 0.0 | 4.2 |
| Husband | 37.8 | 61.7 | 0.5 | 40.9 |
| Jointly with husband | 25.0 | 75.0 | 0.0 | 6.7 |
| Other people | 42.4 | 56.2 | 1.4 | 48.2 |
| **Working status** |  |  |  |  |
| Not working | 51.3 | 48.7 | 0.0 | 29.2 |
| Work throughout the year | 33.2 | 65.1 | 1.7 | 50.6 |
| Work seasonally/Part of the Year | 37.8 | 62.2 | 0.0 | 12.5 |
| Occasionally | 52.7 | 47.3 | 0.0 | 7.7 |
| **Religion** |  |  |  |  |
| Catholic | 50.3 | 49.7 | 0.0 | 18.4 |
| Muslim | 37.0 | 63.0 | 0.0 | 32.7 |
| Anglican | 31.9 | 65.2 | 2.8 | 30.3 |
| Pentecostal | 51.2 | 48.8 | 0.0 | 18.6 |
| **Religion not very important** |  |  |  |  |
| No | 45.9 | 54.1 | 0.1 | 61.3 |
| Yes | 32.5 | 65.4 | 2.1 | 38.7 |
| **Sexual activity** |  |  |  |  |
| Last month | 34.5 | 65.5 | 0.0 | 19.2 |
| Between 2-5 months | 41.6 | 57.2 | 1.2 | 71.8 |
| Between 6-12 month | 75.5 | 24.5 | 0.0 | 6.9 |
| 1 years and above | 13.4 | 84.2 | 2.4 | 2.1 |
| **Parity** |  |  |  |  |
| 0 | 65.4 | 34.6 | 0.0 | 27.1 |
| 1 | 27.1 | 72.9 | 0.0 | 19.5 |
| 2 | 37.9 | 62.1 | 0.0 | 18.2 |
| 3 | 22.5 | 73.1 | 4.4 | 15.0 |
| 4 | 21.1 | 76.7 | 2.2 | 6.9 |
| 5+ | 43.8 | 55.8 | 0.4 | 13.3 |
| Overall | 40.5 | 58.6 | 0.9 | 48.8 |

Findings in Table 2 also show that more women were using long-acting reversible contraceptives (LARC) (59%) compared to short term (40%) and permanent (1%) methods. Short term methods were mainly used by the unmarried (70%), those with secondary/higher education (47%), women in the first quintile (47%), women who make independent decision on FP (70%), Catholics (50%), and those with zero parity (65%).

LARC was mainly used by those living in the mainland slum (63%), married (69%), those with no education/primary level (67%), those in the third wealth quintile (62%), and women who make joint decision with their partners about FP (75%). Further, women who work throughout the year (65%), those whose sexual activity was at least one year (84%), and women with four children (77%) were using LARC to short term methods. Additionally, LARC was also mainly used by those between 30-34 years (78%) while adolescents were using short term methods (74%).

Turning to assessing the distribution of modern family planning use by sexual activity and marital status (Fig 1), the use of mFP among the unmarried and married women who were sexually active within last month preceding the day of interviews were 44% and 58% respectively.

**Fig 1:**
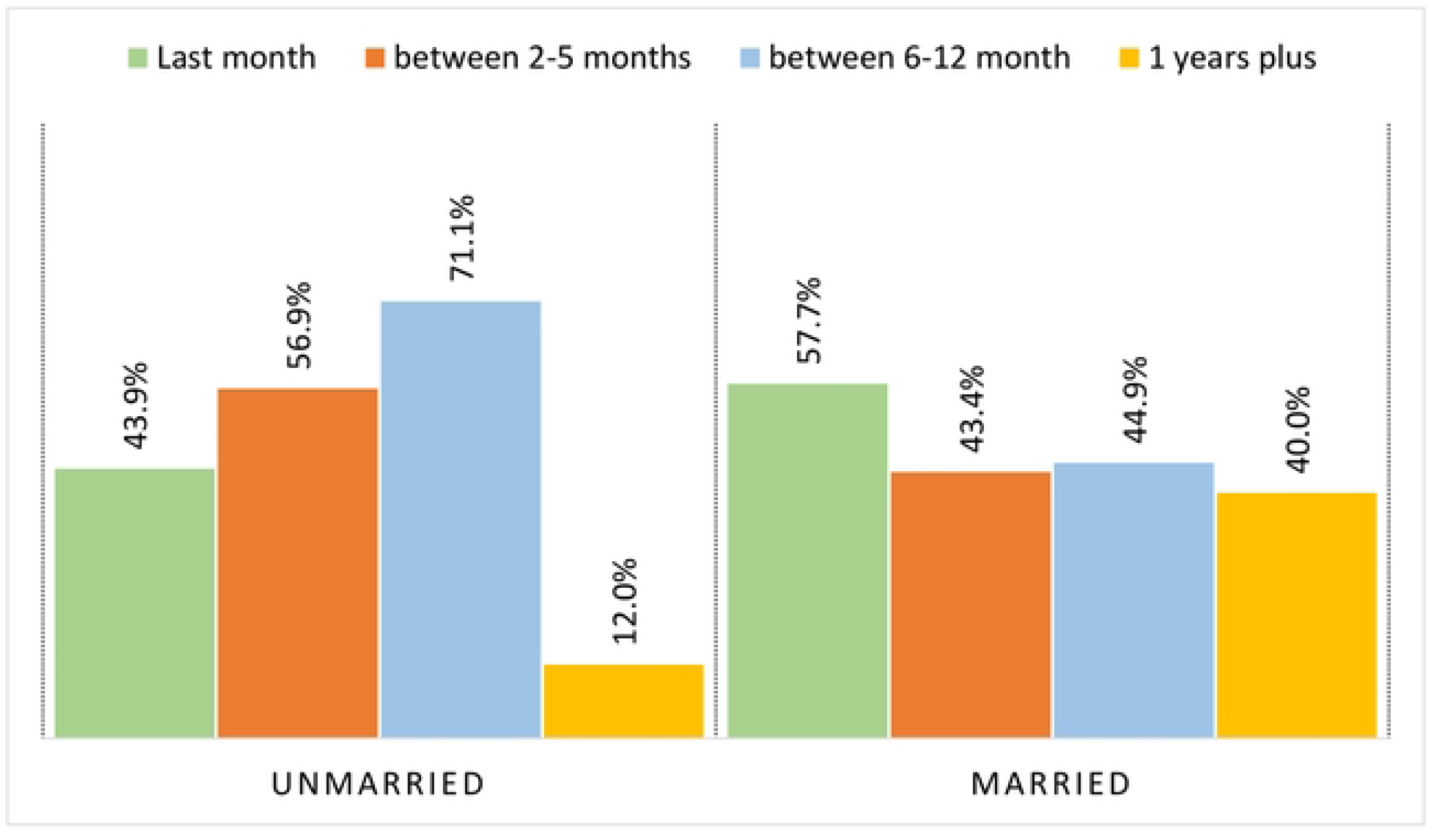
Distribution of modern family planning use by sexual activity intervals and marital status

### Factors associated with modern family planning among women aged 15-49 years

Table 3 presents both the bivariate and multivariate analysis of the factors associated with mFP use. The likelihood of using mFP reduced with women age and increased with parity (Figure 2).

**Fig 2:**
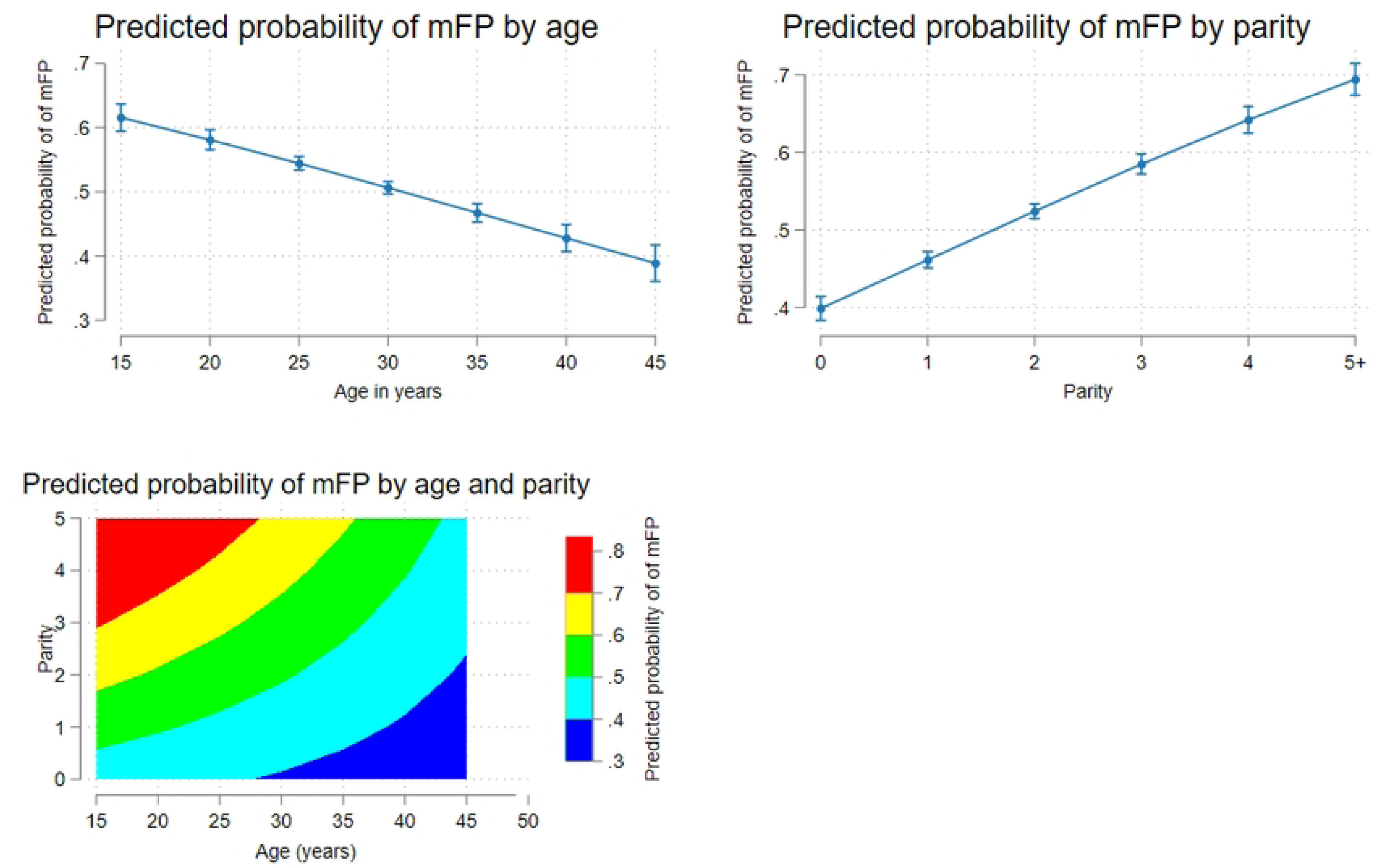
Adjusted probabilities of modern family planning use by age and parity

**Table 3:** Association of the selected variables with modern family planning use

| Variable | OR(95%CI) | Model 1<br>aOR(95%CI) | Model 2<br>aOR(95%CI) |
| --- | --- | --- | --- |
| **Woman's age** | 0.995[0.992-0.999] | 0.976[0.966-0.986]*** | 0.987[0.979-0.996]*** |
| **Parity** | 1.07[1.054-1.085] | 1.404[1.249-1.578]*** | 1.403[1.279-1.54]*** |
| **Age#parity** | - | 0.996[0.992-0.999]* | 0.997[0.994-1.00] |
| **Place of residence** |  | [-] |  |
| Other residential places | [-] | [-] | [-] |
| Landing site slum | 0.926[0.869-0.987] | 0.785[0.573-1.076] | 0.701[0.639-0.769]*** |
| Mainland slum | 1.038[0.983-1.097] | 2.065[1.735-2.458]*** | 1.135[1.053-1.224]*** |
| **Education level** |  | [-] |  |
| No education or primary | 1.196[1.138-1.257] | 1.299[1.047-1.612]* | 1.3[1.181-1.431]*** |
| Secondary/higher | [-] | [-] |  |
| **Wealth index** |  | [-] |  |
| 33.3% of the lowest class | [-] |  |  |
| 33.3% of the middle class | 1.632[1.516-1.757] | 1.832[1.52-2.209]*** | - |
| 33.3% of a better class | 1.442[1.343-1.55] | 5.276[4.082-6.819]*** | - |
| **FP decision making authority** |  | [-] |  |
| Husband | [-] | [-] | [-] |
| Woman alone | 0.521[0.467-0.582]*** | 2.349[1.817-3.037]*** | 3.408[2.701-4.301]*** |
| Jointly with husband | 1.052[0.949- 1.167 ] | 0.839[0.707-0.996]* | 0.986[0.854-1.137] |
| Other people | 0.841[0.800-0.886]*** | 0.555[0.499-0.618]*** | 0.753[0.691-0.82]*** |
| **Working option** |  | [-] |  |
| Not working | [-] | [-] | [-] |
| Work throughout the year | 1.441[1.365-1.52] | 1.152[1.056-1.257]*** | 1.26[1.17-1.356]*** |
| Work seasonally/Part of the Year | 1.607[1.48-1.744] | 1.369[1.195-1.569]*** | 1.667[1.48-1.879]*** |
| Occasionally | 2.253[2.023-2.509] | 0.991[0.839-1.17] | 1.631[1.422-1.872]*** |
| **Religion** |  | [-] |  |
| Catholic | [-] | [-] | [-] |
| Muslim | 0.909[0.847-0.975] | 0.827[0.743-0.921]*** | 0.894[0.814-0.982]* |
| Anglican | 1.047[0.974-1.125] | 1.076[0.964-1.2] | 1.023[0.93-1.126] |
| Pentecostal | 0.732[0.678-0.791] | 1.192[1.057-1.344]** | 1.086[0.982-1.201] |
| **Religion not very important** |  | [-] |  |
| No | [-] | [-] |  |
| Yes | 1.036[0.986-1.088] | 1.217[1.125-1.316]*** | 1.202[1.124-1.286]*** |
| **Recent sexual intercourse** |  | [-] |  |
| Within last month | [-] | [-] | [-] |
| between 2-5 months | 0.644[0.602-0.69] | 0.634[0.58-0.693]*** | 0.602[0.56-0.648]*** |
| between 6-12 month | 0.832[0.757-0.916] | 0.744[0.664-0.834]*** | 0.915[0.827-1.012] |
| 1 years plus | 0.114[0.099-0.13] | 0.154[0.128-0.184]*** | 0.128[0.11-0.149]*** |
| **Level of education# FP decision making authority** |  | [-] |  |
| No education or primary level#Woman alone | - | 0.241[0.127-0.457]*** | 0.075[0.044-0.13]*** |
| No education or primary level#Jointly with husband | - | 0.598[0.441-0.809]*** | 0.645[0.498-0.836]*** |
| No education or primary level#Other people | - | 1.286[1.09-1.517]** | 0.937[0.816-1.075] |
| **Place of residence#wealth index** |  | [-] |  |
| Landing site slum#33.3% of middle class | - | 1.265[0.88-1.816] | - |
| Landing site slum#33.3% of better class | - | 0.182[0.123-0.267]*** | - |
| Mainland slum#33.3% of middle class | - | 0.421[0.337-0.526]*** | - |
| Mainland slum#33.3% of better class | - | 0.145[0.11-0.192]*** | - |
| **Wealth index#Level of education** | - | [-] | - |
| 33.3% of middle class #No education or primary level | - | 0.917[0.728-1.156] | - |
| 33.3% of better class#No education or primary level | - | 1.119[0.891-1.406] | - |

\*\* $p < 0.01$ , \* $p < 0.05$ ; OR is the unadjusted odds ratio; aOR is the adjusted odds ratio at 95% confidence interval

An additional year would lead to a 2% reduction in mFP, while an additional birth would lead to a 40% increase in mFP use (Table 3). The interaction between age and parity indicates that the likelihood of mFP was higher among women within the youthful age (20-30) with at least 3 births (**Fig 2**. Compared to women in non-slum mainland areas, women residing in mainland slums were twofold more likely to use mFP. Changing the base to landing site slum residence (Table 4), women residing in mainland slums where almost threefold more likely to use mFP. Overall, the likelihood of using mFP was lower among women residing in landing site slum (Fig 3).

**Fig 3:**
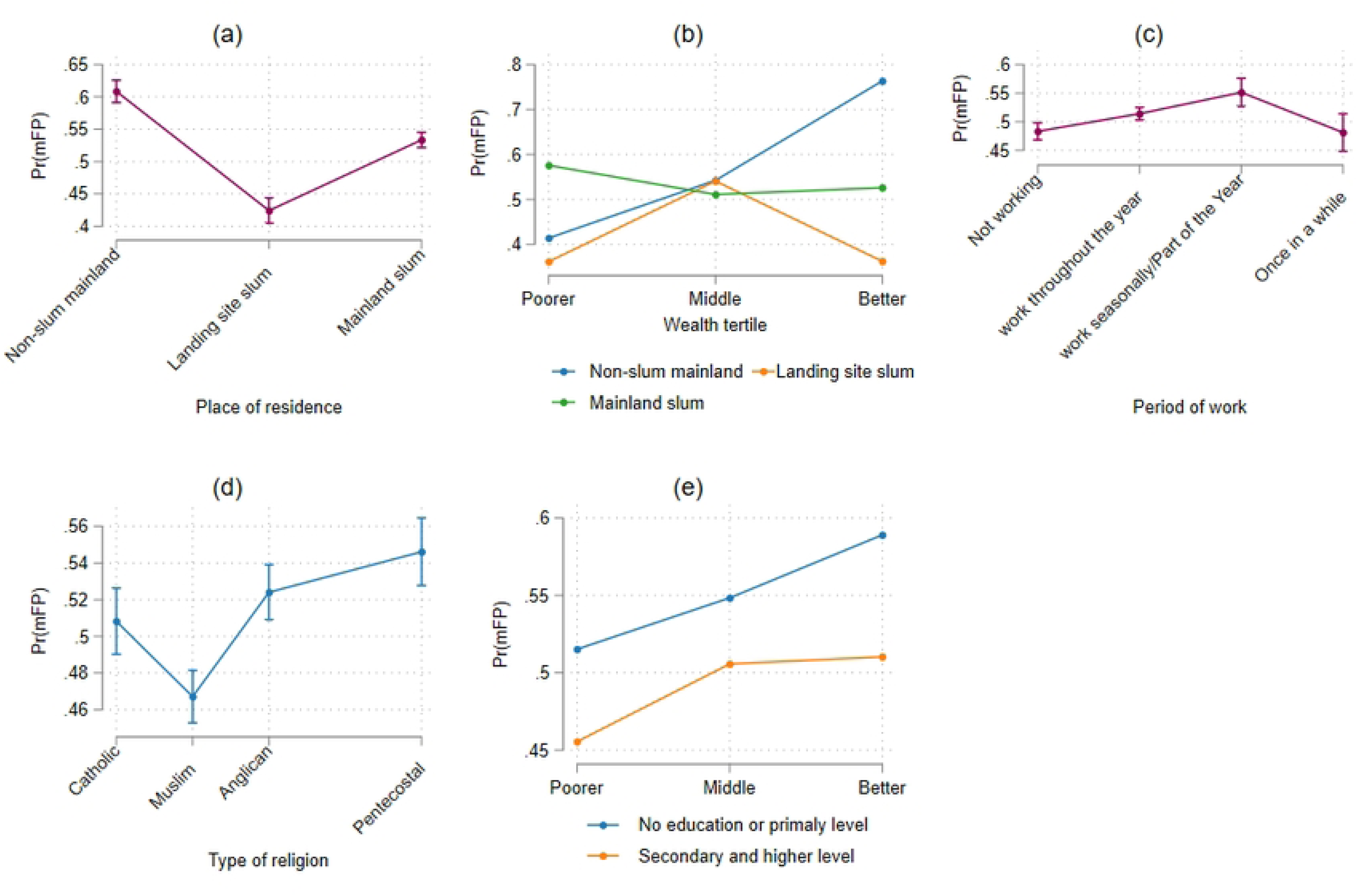
Predictive probability of mFP by (a) place of residence, (b) wealth index and place of residence, (c) work type, (d) religion, (e) wealth and education level

**Table 4:**
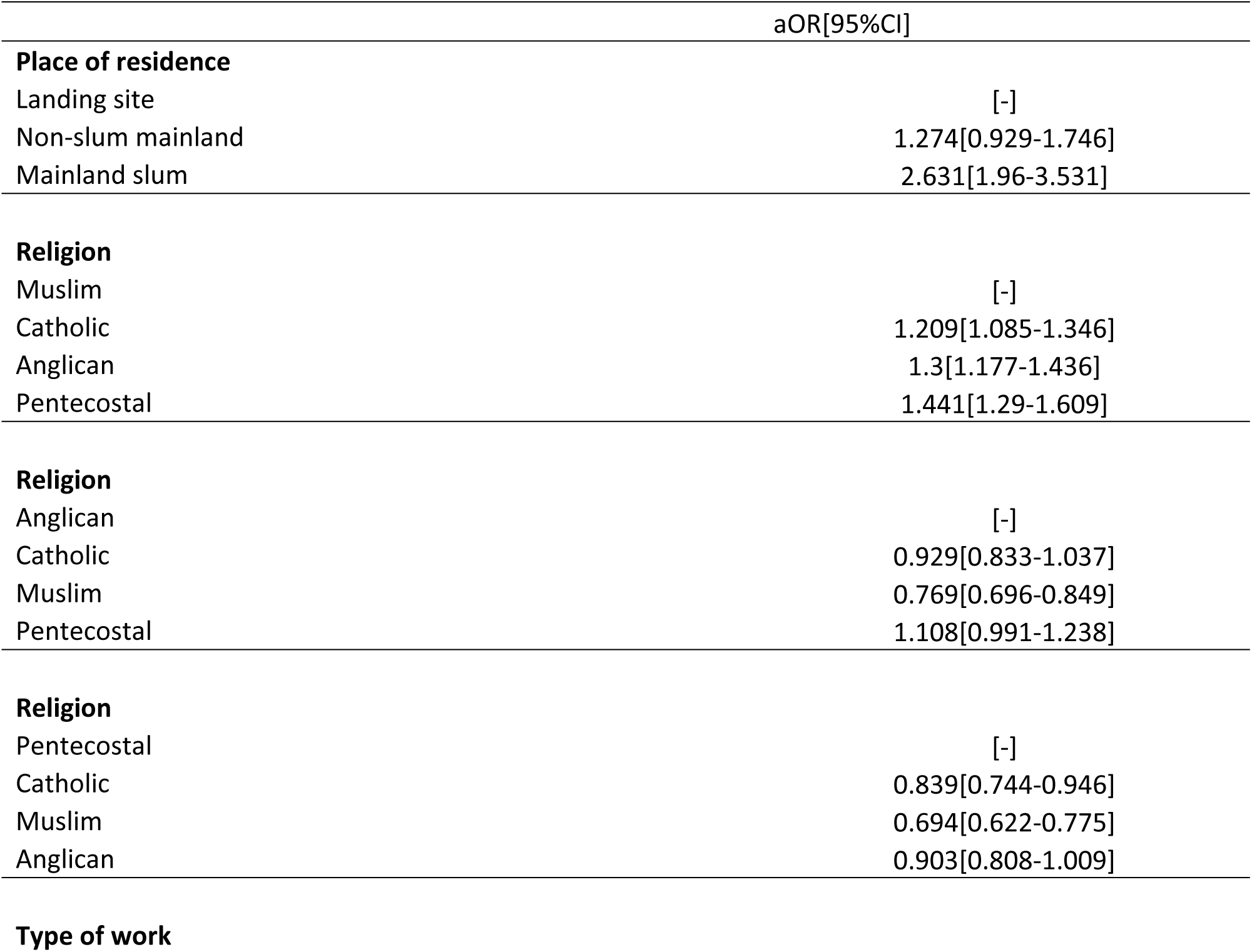
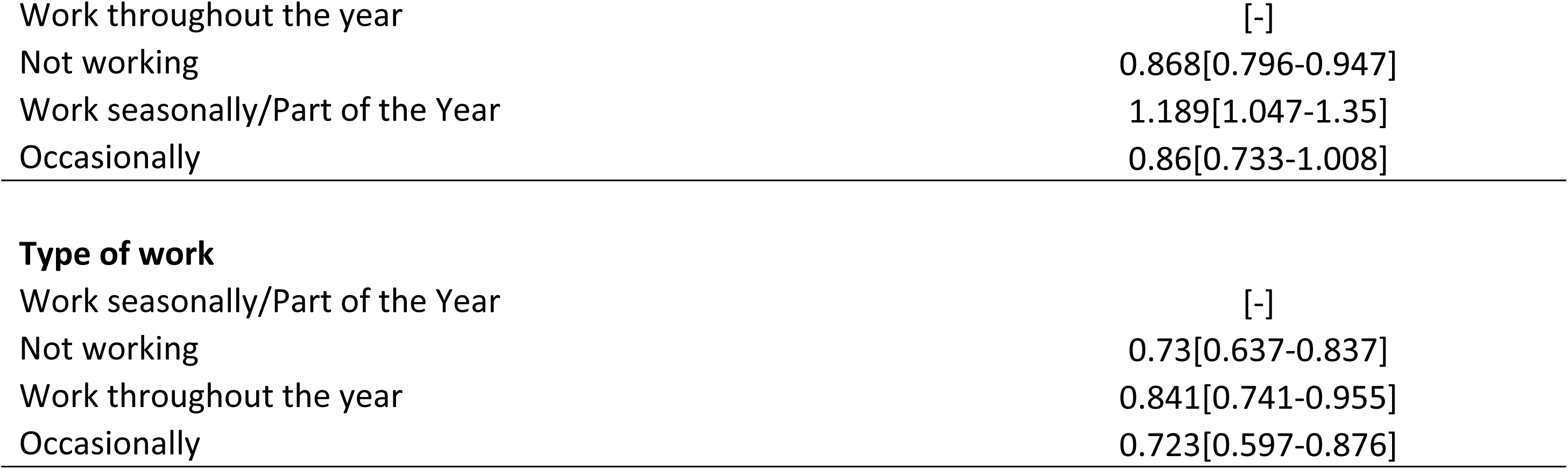
Changes in variable base using Model 1 in Table 3

|  | aOR[95%CI] |
| --- | --- |
| **Place of residence** |  |
| Landing site | [-] |
| Non-slum mainland | 1.274[0.929-1.746] |
| Mainland slum | 2.631[1.96-3.531] |
| **Religion** |  |
| Muslim | [-] |
| Catholic | 1.209[1.085-1.346] |
| Anglican | 1.3[1.177-1.436] |
| Pentecostal | 1.441[1.29-1.609] |
| **Religion** |  |
| Anglican | [-] |
| Catholic | 0.929[0.833-1.037] |
| Muslim | 0.769[0.696-0.849] |
| Pentecostal | 1.108[0.991-1.238] |
| **Religion** |  |
| Pentecostal | [-] |
| Catholic | 0.839[0.744-0.946] |
| Muslim | 0.694[0.622-0.775] |
| Anglican | 0.903[0.808-1.009] |
| **Type of work** |  |
| Work throughout the year | [-] |
| Not working | 0.868[0.796-0.947] |
| Work seasonally/Part of the Year | 1.189[1.047-1.35] |
| Occasionally | 0.86[0.733-1.008] |
| **Type of work** |  |
| Work seasonally/Part of the Year | [-] |
| Not working | 0.73[0.637-0.837] |
| Work throughout the year | 0.841[0.741-0.955] |
| Occasionally | 0.723[0.597-0.876] |

Whereas the odds of using mFP increased with the wealth status (Middle: aOR=1.832, 95%CI=[1.52-2.209] and Better: aOR=5.276, 95%CI=[4.082-6.819]), introducing an interaction between the place of residence and wealth tertile highlight that only women in poor household position in non-slum mainland areas were less likely to use family than those in the middle and better wealth household position (Fig 3). Women of poor household wealth position in mainland slum were more likely to use mFP, while those in poor and better household wealth position residing in slums around the landing sites were less likely to use mFP. In addition, compared to those with secondary or higher education level, women with no level or primary education were 29% more likely to use mFP. Introducing an interaction between wealth and education indicated that the likelihood of mFP use among women with no or low levels of education increased with their wealth position and estimates remained higher than the educated one (Fig 3).

Turning to the religion, the odds of using mFP was 23% higher among those who considered religion as not most important in their daily life. Relative to Catholics, the odds of using 17% lower among the Muslim and 19% higher among Pentecostal. Changing the base to Muslim affiliation (Table 4), the odds of using mFP were 20% among Catholic affiliates, 30% among Anglicans, and 44% among Pentecostal affiliates. Similarly, changing the base to Anglican affiliation (Table 4), the odds were 23% lower among the Muslim affiliates. Finally, changing the base to Pentecostal affiliation (Table 4), the odds were 17%, 31%, and 10% lower among the Catholics, Muslim, and Anglican, respectively. Overall, the likelihood of mFP use is lower among Muslim and Catholic affiliates (Fig 3).

Considering the level of employment, relative to women who were not working, the odds of using mFP were 15% and 37% higher among those working throughout the year and those working seasonally or parttime in a year, respectively (Table 3). Changing the base to working throughout the year (Table 4), the odds of using mFP were 24% lower and 18% higher among women who were not working at all and those working seasonally, respectively. Changing the base to seasonal working (Table 4), the odds of using mFP were 27%, 16%, and 28% lower among women who were not working, those who were working throughout the year, and those who were working occasionally. Overall, the likelihood of using mFP were lower among those who were not working and those who were working occasionally (Fig 3).

While at bivariate the odds of using mFP among women whose decision to use mFP independent of their husbands or social networks were low, we observed a change in the sign at multivariate: the odds of using mFP were twofold among women whose decision to use mFP were independent of husband or other social networks. When we introduced an interaction between the women decisions authority and education level, the likelihood of using mFP was higher among educated women whose decisions to use mFP were independent of husband or social network (Fig 4).

**Fig 4:**
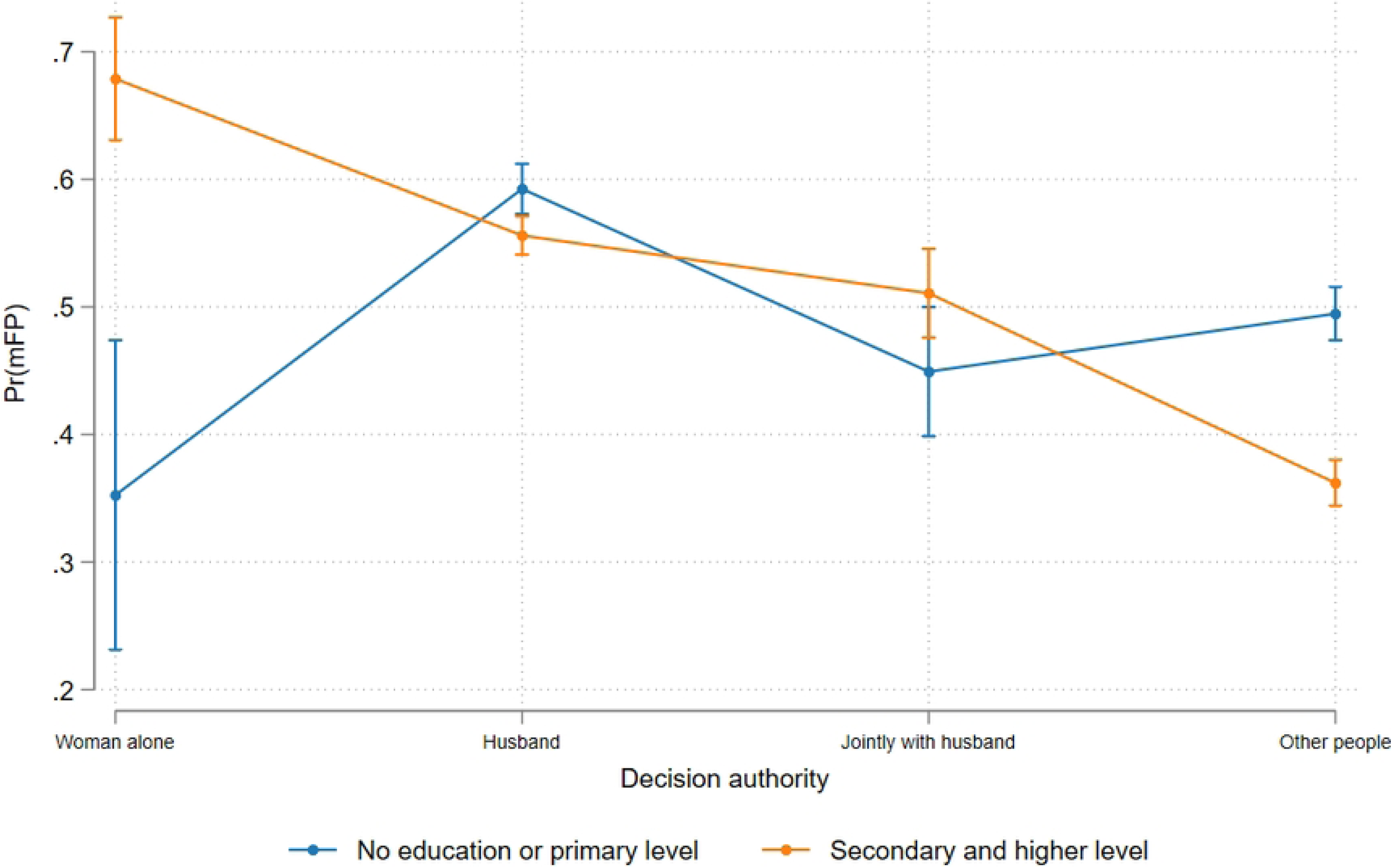
Predictive probability of mFP decision making authority and education

Table 5 shows the inequality in mFP use. Overall, 24% of heterogeneity in mFP use was due to the factors included in the model. The total inequality due to place of residence, socio-economic status, demographic factors, and family planning decisions authority was 18%.

**Table 5:** Inequality in modern family planning use by individual factors

| Variable | Value | Percentage |
| --- | --- | --- |
| Group 1: Wealth index | 0.0085 | 3.6% |
| Group 2: Education level | 0.0166 | 7.1% |
| Group 3: Employment type | 0.0039 | 1.7% |
| Group 4: Religion type and its importance | 0.0248 | 10.5% |
| Group 5: Period of recent sexual intercourse | 0.0598 | 25.4% |
| Group 6: Age and parity | 0.0392 | 16.7% |
| Group 7: Authority over family planning use | 0.0398 | 16.9% |
| Group 8: Urban place of residence | 0.0428 | 18.2% |
| **TOTAL** | **0.2353** | **100.0%** |

## Discussion

In this study, we assessed individual factors that affect modern family planning (mFP) use in emerging towns of Uganda. In this study, mFP was estimated at 48%, with close to 60% using long-acting reversible methods. While the prevalence of mFP use was above the national estimates, we identified inequalities in family planning use. Overall, the factors that we considered in the model explained 24% of the inequality in mFP use, with demographic factors, socio-economic measures, place of residence and family level authority contributing the largest share. 76% of the inequality of opportunity in mFP use that was due to unexplained circumstances could be as a result of supply side factors and some of the demand side factors [18] that were not captured in this study. Some of the supply side factors may include the availability of family planning services in the health facilities including trained health workers, and health facility-community distance [18]. The demand side factors may include the social and cultural issues and women agency/autonomy in accessing the services. The individual factors that explain the 24% of variation in mFP use were women employment, religion affiliation, women autonomy in FP use, age, parity, education level, household wealth position, recent time of sexual intercourse, and urban place of residence (slum status).

Consistent with some studies [19, 20], women who were working were more likely to use family planning compared to those who were not working. In this study, we found that the use of Long-Acting Reversible Contraception was high among women who work throughout the year. However, the relationship between employment and family planning use could be complex to understand. First, to our understanding, women who are engaged in any income generating activity usually fear to lose their source of income in case of becoming pregnant and the conflicting family and employment burden. Secondly, one would also argue that women engaged in fulltime employment have a burden of balancing the family and employment activities and thus have limited time to access reproductive health services including family planning [21]. Some scholars have noted how the type of employment is critical in determining access to family planning services [21, 22]. For instance, the variation in which the different jobs encourage autonomy and their extent to accommodate flexibility in scheduling work activities creates differences in healthcare seeking including FP access [21]. Lastly, women who are not working have less autonomy, which affects their agency in accessing family planning services. We recommend a comprehensive study that seeks to understand family planning use and employment dynamics.

We observed religion affiliation differentials in mFP use. We found that women who considered religion not to be very important in their daily life were more likely to use mFP. Religion is well known as one of the most important determinants of population (believer’s) behaviors, including healthcare seeking [23–25]. In some religions, birth control through modern FP use is taken to contradict their religious beliefs. Religious beliefs on issues of birth control and modern family planning methods adoption can differ greatly among Aglican, Catholics, Muslims, and emerging Pentecostal churches [23, 26]. For instance, whereas traditional Protestant churches may allow mFP, Catholic Church allows only natural methods of contraception and prohibit modern birth control methods. Muslim and traditional churches (Anglican and Catholics) encourage many children as gift from God, which limits the use of the family planning. There is a need to design interventions that engage religious leaders as potential change agents for promoting FP use. Fortunately, interventions that have involved religious leaders in FP advocacy based on the biblical and Quran text have provided promising results [23, 27].

Women whose decision to use mFP was independent of their husband and other social network members were more likely to use mFP. Nonetheless, women’s autonomy to use mFP was mediated by the level of education. In this study, women with secondary or higher level of education and whose decision to use mFP were independent of other authorities were more likely to use mFP. On the other hand, the less educated whose decision to use mFP depended on their husband and other social networks were less likely to use mFP. In sub-Saharan Africa, many decisions on sexual and reproductive health are scrutinized by partners and older female relatives, who generally exert more influence on women’s use of health services [20]. This finding suggests the need to design gender empowerment interventions that target both men and women in the community.

The results show that respondent’s age was associated with mFP. Older women were less likely to use mFP compared to the younger ones. However, among those aged 20 years and above, at least 50% of the mFP users were using a long-acting reversible methods. Furthermore, women with parity of 5+ were more likely to used mFP, which highlights an opportunity for women who would either want to space or limit the number of children. The use of long-acting reversible FP methods was found to progressively increase with parity and was highly used among those with 4^th^ parity and above. When we introduced the interaction between age and parity, we found that women within the youthful age (15-30) with at least 3 births where more likely to use mFP. Additionally, the odds of mFP were observed to regressively reduce with the increase in recent period of sexual intercourse. Indeed, in the absence of contraceptive use, sexual activity is an important factor that is positively associated with the risk of pregnancy [28] and therefore understanding how the recent sexual intercourse are associated with mFP use give us insight into the use of mFP among sexually active people [28]. Close to 4 out of 10 unmarried women and 6 out of 10 married women who had sex in the last month prior to the day of data collection used mFP, which presents an opportunity for improved demand for mFP use. Such finding gives insight into community demand for controlling fertility and thus suggest the need for availing FP method mix, particularly the long-term methods.

The odds of using mFP were also observed to progressively increase with wealth quintile, being 80% higher and 5 times higher among the middle and better compared to the poorest. Enormous studies done in sub-Saharan Africa have found a strong positive effect of wealth index on mFP use [29–36]. Such findings show how financial constraint could have role in reducing mFP usage among poor women. The financial constraints may include the dyad of having money to purchase the appropriate method and to pay for transport to reach the health facilities. Interventions that ensure the availability of mFP at minimal or no cost could therefore improve usage.

Our study provides surprising results. Inconsistent with other studies in the same setting [19, 20, 37], women with secondary education or higher were less likely to use mFP compared to those with no formal education or primary education. When we introduced the interaction between education and wealth, the rich who were not educated were more likely to use mFP than their educated counterparts. Furthermore, we also found that women residing in the mainland and landing site slum areas were more likely to use modern family planning services than those in mainland non-slum places of residences. Residential variation in mFP use is influenced by various factors including the availability of and distance to service providers. From our assessment, we found that many small clinics are concentrated in small towns within the mainland. Also, we noted that the mainland slums are close to the urban business centre and can easily accesses the services, while the mainland non-slum residential areas are apart from the urban business centres, and therefore, use of health services including family planning may be hard for those with financial constraints. The findings challenge the usually dichotomous categorisation of considering only slum dwellers as the most deprived without considering the other marginalised population that are living within the rich neighbourhood. These findings showcase the dynamics in utilizing health services within an urban context and thus suggesting the need for interventions that target all places of residences within the urban setting.

## Strengths and limitations

The strength of this study is that it uses a large pool of dataset to explain the dynamics in the inequality of modern family planning use in emerging urban areas of Uganda. The limitations of the study result from the nature of cross-sectional studies. First, our analysis approach does not establish a direct cause–effect relationship though our understanding of the contribution of each observed factors provide an insight into the community and individual FP utilization determinants that should be considered in programming and decision-making. Secondly, since the data is based on respondents’ self-reported information, some sexual and fertility related questions could have been answered to please the interviewer, which could have led to social desirability bias. Nonetheless, during data collection we assured respondents’ confidentiality and anonymity steps. Lastly, some of the demand side variables such as the social and cultural issues factors, and supply side factors such as availability of services were not capture, which would explain why the heterogeneity in mFP use was largely (66%) due to unexplained factors. Nonetheless, most of the factors that we included have been well studied as the measures of inequality and women empowerment.

## Conclusion

Our study is the first to provide literature on the dynamics in inequalities of modern family planning use in an urban setting of low-income countries. While the urban areas are expected to have better utilization of family planning services, our findings highlight the intra-urban inequality in mFP use that are due to socio-economic and demographic factors, place of residence and family planning decisions authority. The findings highlight the need for intervention that are tailored to the different groups of urban residents. For instance, the package of interventions should consider the places of work and places of residence regardless of socioeconomic status. However, strengthening the supply-side to be able to meet the demand is a sufficient condition that should be implemented alongside demand-side interventions. Since most of the factors that we identified to influence mFP use such as employment status, education level, wealth status are measures of women agency and empowerment, we suggest for demand-side and supply-side interventions that strengthen the autonomy of women in family planning use decision.

## Data Availability

The Urban Thrive Project data are not shared publicly but can be made available to other researchers with approval by the project research investigators. Requests for data access may be sent to Rornald Muhumuza Kananura at.

